# *“… I tried to commit suicide…”:* Understanding the intersections between mental health, HIV and teenage pregnancy

**DOI:** 10.1101/2025.04.02.25325150

**Authors:** Hanani Tabana, Takunda Satumba, Diane Cooper, Martina Lembani

## Abstract

**Background:** Mental health is an essential component of well-being, yet marginalized groups, particularly adolescents, face systemic inequities and barriers to accessing care. This paper describes the experiences of adolescent girls in their access to healthcare services and how these experiences might have contributed to their mental health and well-being in the Western Cape province, South Africa.

**Methods:** This qualitative study employed narrative and semi-structured interviews to explore the sexual and reproductive health and mental health well-being of adolescent girls aged 15–19. Participants included adolescent girls in various categories; pregnant, postpartum, HIV positive, and HIV negative, recruited from three youth-friendly primary healthcare facilities using purposive and snowball sampling methods. A total of 17 adolescents, 4 healthcare providers, and 4 parents were interviewed and a focus group was held, involving 6 sub-district frontline healthcare program managers.

**Results:** The factors contributing to mental health among the adolescents were broadly categorised under five themes, 1) Navigating the impact of unintended pregnancy, 2) Negotiating the home environment and other relationships, 3) Barriers to access to services at facility level, 4) Social challenges, and 5) Improving mental health services.

**Conclusion:** This study explored factors that contribute to or hinder the mental well-being of adolescent girls and the barriers to accessing mental health services. Designing tailored approaches to the identified factors and systemic challenges that counter mental distress for this age group can significantly mitigate the impact on their mental health.

## Introduction

Mental health is increasingly recognized as a critical component of overall well-being (Gautam, Jain, Chaudhary, Gautam, Gaur, and Grover, 2024). Yet, disparities in access to mental healthcare persist globally (Shisana, Stein, Zungu, and Wolvaardt, 2024). Marginalised populations, including adolescents, experience heightened barriers to mental health services due to systemic inequities, stigma, and socioeconomic challenges (World Health Organization, 2024). Globally, one in seven adolescents between the ages of 10 and 19 experiences a mental disorder; however, their access to optimal mental healthcare services is inadequate (WHO, 2024). In sub-Saharan Africa, adolescents face inequitable access to adequate mental health services compounded by poverty, limited healthcare infrastructure, bullying in schools exacerbated by social media, family dysfunctions, difficult home environments, and unsafe neighbourhoods (Herman, Stein, Seedat, Heeringa, Moomal and Williams, 2016; Mokitimi, Jonas, Schneider, and De Vries, 2019; Källmén and Hallgren, 2021; Shisana et al., 2024).

In Southern Africa, teenage girls who are pregnant and/or living with HIV represent one of the most vulnerable and underserved groups (Toska, Laurenzi, Roberts, Cluver, and Sherr, 2020). Studies highlight that South Africa, with one of the highest HIV prevalence rates globally, faces an adolescent pregnancy rate of approximately 16% and a significant burden of mental health conditions among adolescent girls and young women (Statistics South Africa, 2022; Shisana et al., 2024). Mental health issues, including depression and anxiety, are disproportionately prevalent among pregnant teenagers, particularly those living with HIV due to intersections of stigma, social exclusion, and health challenges (Mkhize, van der Westhuizen, and Sorsdahl, 2024; Cluver, Orkin, Yakubovich and Sherr, 2020). Despite their high burden, adolescents often continue to experience barriers to sufficient mental healthcare services and facilities (Makwero, Mahenge, and Smith, 2019).

In South Africa, mental health services for pregnant teenagers are critically inadequate, and services are underfunded and under-resourced, limiting their access to vulnerable adolescents (Mokitimi et al., 2019; Shisana et al., 2024). Ensuring equity in mental healthcare access for pregnant teenagers living with HIV is crucial, aligning with global commitments to achieving universal health coverage and the Sustainable Development Goals (SDGs), particularly those aimed at reducing health disparities (WHO, 2022). This article seeks to explore mental health challenges experienced by adolescents and to understand their access to mental health services within the broader South African sexual and reproductive health services context. These findings aim to address a knowledge gap on adolescent mental health and their access to mental health services, based on insights shared by teenagers, caregivers, and healthcare providers.

### The Socio-Ecological Model

Various frameworks have been used to conceptualise access to healthcare, including patient-centered approaches (Levesque, Harris, and Rusell, 2013). The socio-ecological model developed by Bronfenbrenner in the late 1970s (McLeroy, Bibeau, Steckler, and Glanz, 1988) provides a comprehensive framework for understanding the complex factors affecting healthcare. This framework conceptualizes individuals as nested within multiple levels of influence, including intrapersonal, interpersonal, organizational, community, and policy levels. It has been extensively adopted by various researchers to explain various phenomena. We hypothesize that adolescent mental health and access to sexual and reproductive care services are impacted by various factors and at different levels, as depicted in Fig 1. This model had been adopted from an ecological model for health promotion by McLeroy et al. (1988).

**Figure 1:**
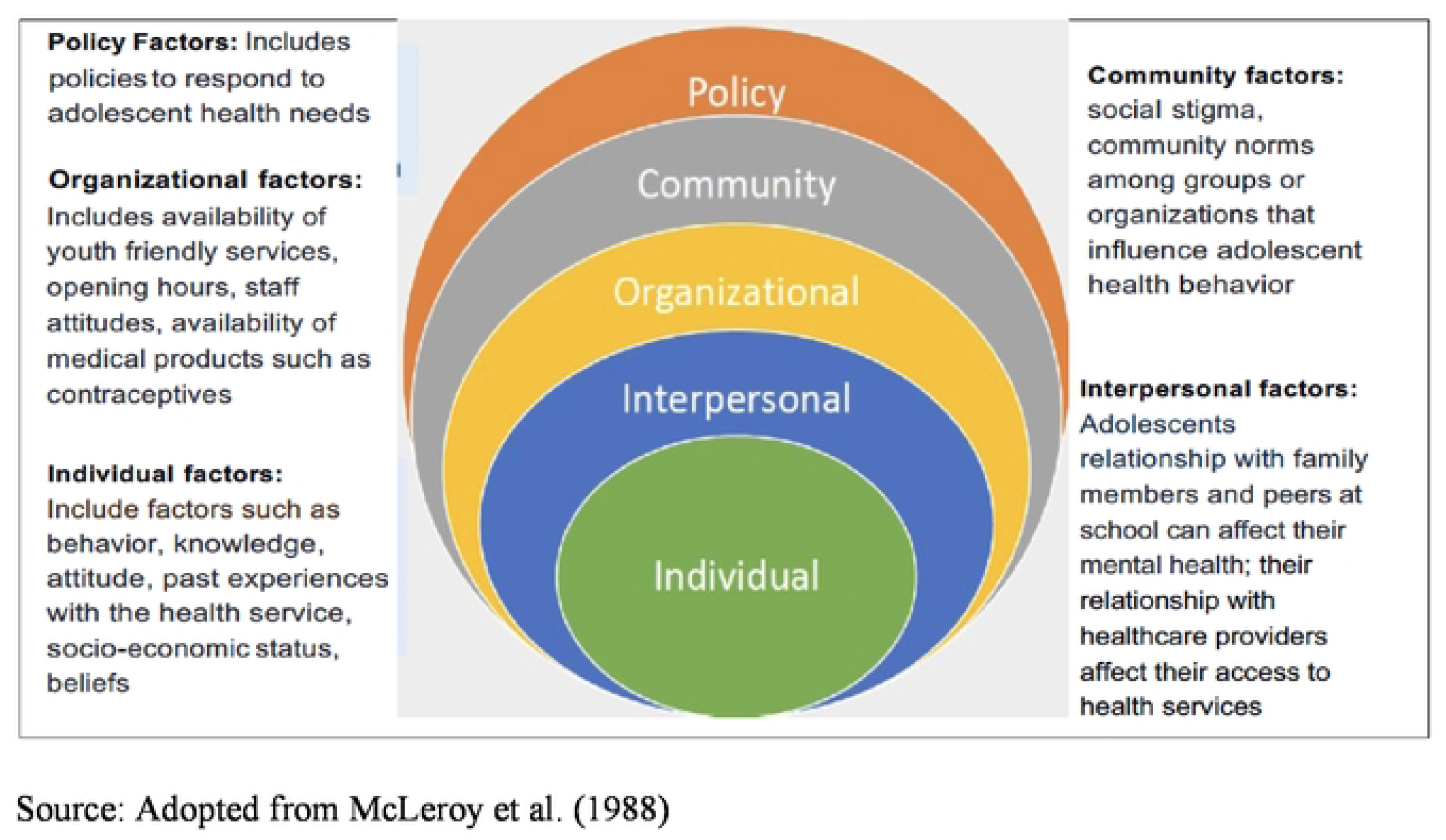
An Ecological Model for Health Promotion

The study used this framework to identify key factors that may influence the mental health and well-being of adolescent girls as well as their access to health services in the City of Cape Town.

## Materials and Methods

### Study design

This study used a qualitative approach that involved narrative interviews, semi-structured interviews, and a Focus Group Discussion (FGDs) with selected participants in the City of Cape Town.

### Study population

Participants for the study included adolescent girls aged 15-19 years in Cape Town. The adolescent girls comprised different categories: those who were pregnant, those who were postpartum, those who had never been pregnant, those living with HIV, those who were not living with HIV, as well as those who had overlapping characteristics i.e. pregnant or postpartum and HIV positive or negative. In addition, healthcare providers who worked in youth-friendly clinics and parents/caregivers of adolescent girls were selected to participate in the study. Finally, a focus group discussion was held with a purposively selected sub-district team, which included frontline managers.

### Sampling

Purposive sampling was used to select adolescent girls who met the following study inclusion criteria:

1. Between the ages of 15 and 19 years
2. Pregnant and not living with HIV
3. Pregnant and living with HIV
4. Not pregnant and not living with HIV
5. Postpartum and living with HIV
6. Postpartum and not living with HIV

This was done to gather different life stories and be able to compare narratives across groups. Adolescents were identified through Primary Health Care facilities in Cape Town after obtaining approval from the National Department of Health Research Committee and permission from the PHC facility managers. Permission was granted for 5 selected youth-friendly health facilities. However, adolescents were recruited from only three facilities based on the availability of participants meeting the participation criteria and willing to participate in the study. The interviewer recruited adolescents attending various services in the health facilities. Healthcare providers assisted with the identification of eligible participants, who were then approached by the interviewer in the waiting room. Those willing to be interviewed were privately interviewed within the health facility. Snowballing was also used to identify additional adolescent participants in the study. Healthcare providers involved in the provision of youth-friendly services at the identified health facilities were purposively selected for interviews. Parents/caregivers of the adolescents were requested for interviews and those willing to be interviewed were telephonically interviewed. Attempts were made to interview partners of the pregnant and postpartum adolescents, however, they declined to participate in the study. A total of 17 interviews were held with adolescent girls, 4 with parents, and 4 with healthcare providers. Additionally, 6 sub-district frontline healthcare program managers participated in a focus group discussion.

### Data collection process

The interviewer visited the health facility and introduced the research topic to the facility managers, who then delegated one healthcare worker to assist with identifying prospective participants. Eligible participants were approached and introduced to the study objectives and were invited to participate in the study. For those who consented to participate, arrangements were made for a private interview in a room at the clinic.

The interviews comprised narratives of the adolescents’ childhood experiences in general and their experiences with being pregnant, postpartum, or HIV positive, family environment, and how their specific circumstance affected their mental well-being. The adolescents were asked about access to health services when seeking sexual and reproductive health services at the health facilities and awareness of mental health policies.

Data was collected between July and October 2021. The interviews ranged between 30 minutes and an hour. The interviews were recorded with the participants’ permission.

### Data analysis

A thematic analysis approach was used to analyse data. A deductive thematic approach was used based on the socio-ecological framework presented in Figure 1. First, all the interview recordings were transcribed verbatim. The transcripts were read several times by the researchers to identify key issues and were coded. The researchers discussed the identified codes. Themes and subthemes were placed under each of the SEM domains as presented in the socio-ecological model. Themes and subthemes were identified inductively. To ensure the objectivity of the data, four researchers (HT, TS, DC, and ML) coded the data separately, discussed the identified codes, and reached a consensus. Members included reflexivity when analysing data to deter biases in interpreting the data.

## Results

Five broad themes emerged from the data analysis, as presented in Table 1. These include: 1) navigating the impact of unintended pregnancy, 2) negotiating the home environment and other relationships, 3) barriers to accessing services at the facility level, 4) social challenges, and 5) barriers to accessing mental health services. Using the Bronfen Benner socio-ecological model, themes were categorised into: individual, interpersonal, organisational, community, and policy-level domains. A summary of these themes and their corresponding subthemes is presented in Table 1.

**Table 1:**
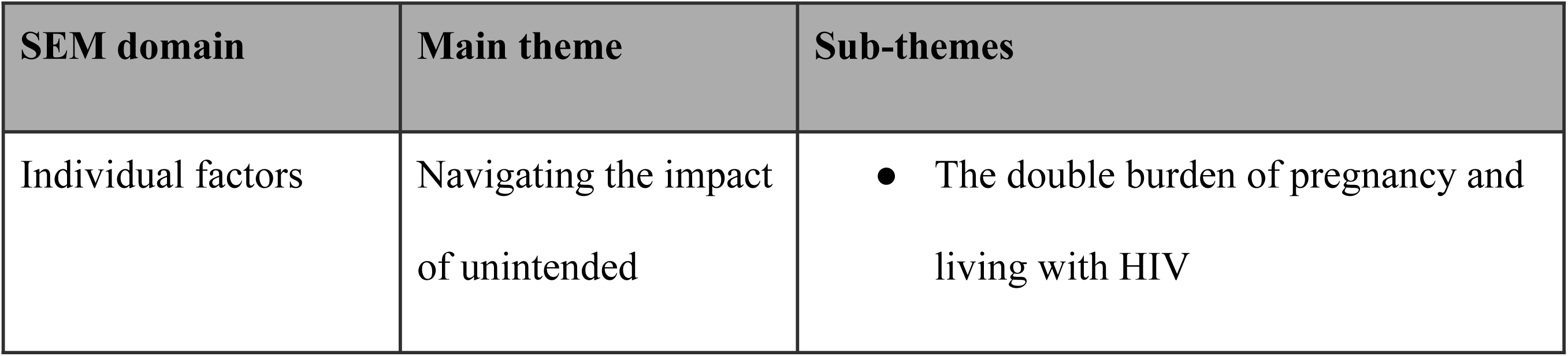
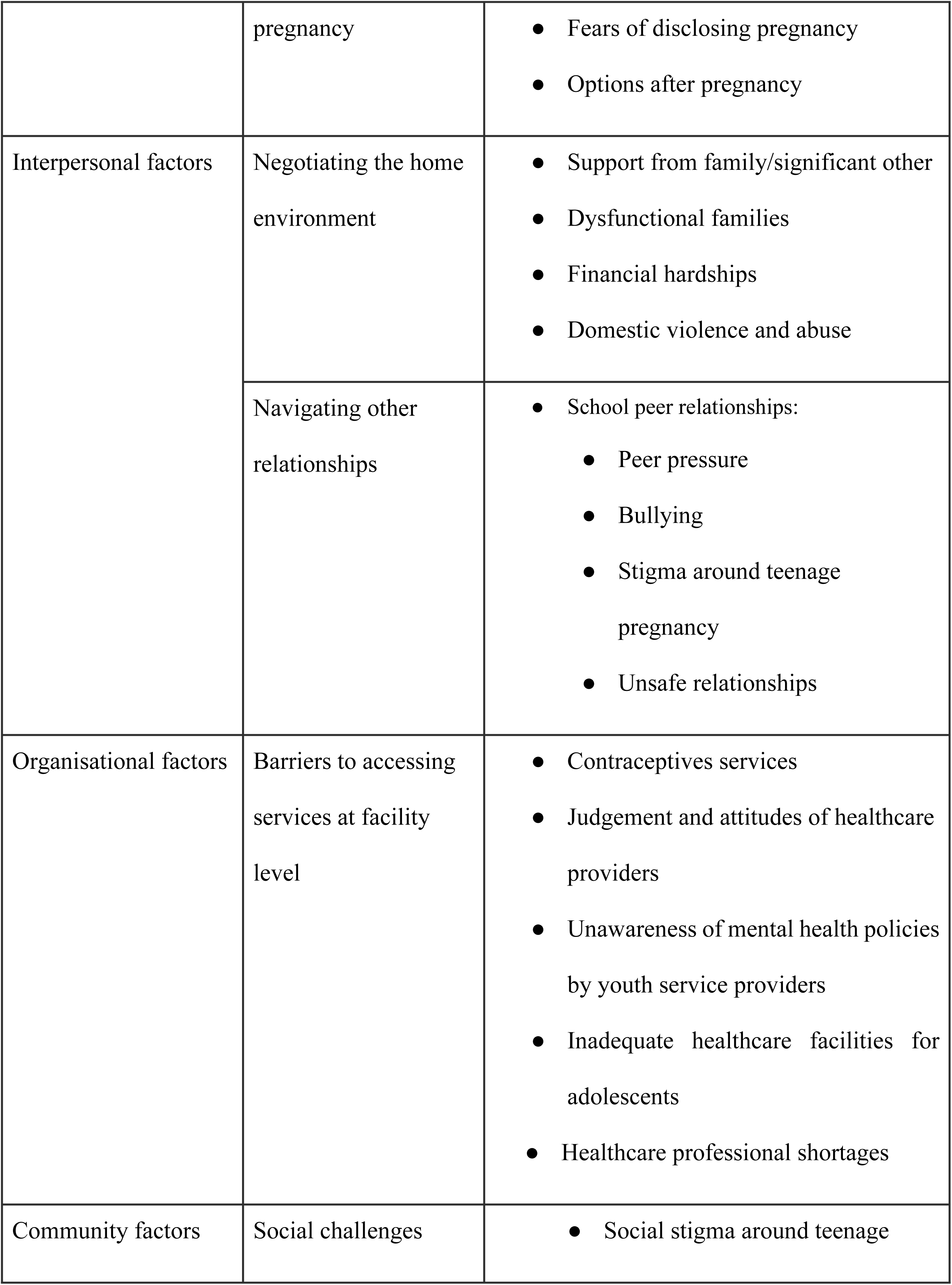
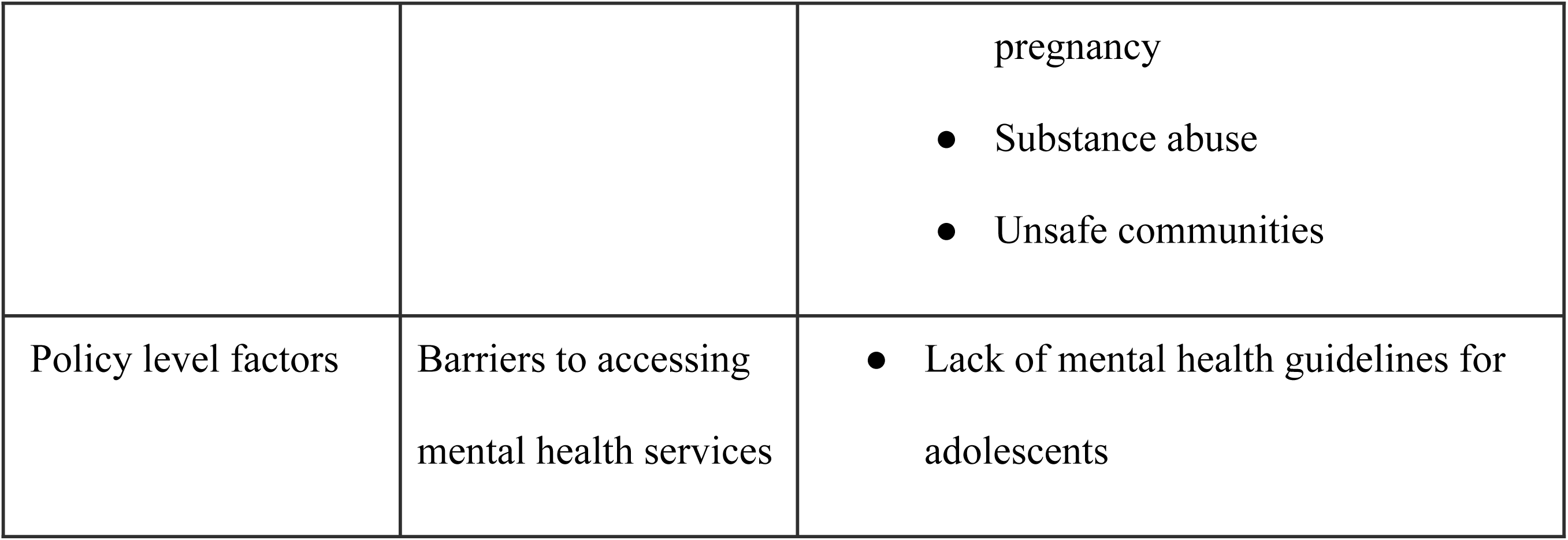
Summary of sub-themes under each domain of the SEM.

Table 1 summarizes the key themes and subthemes/factors that were identified under each level of the socio-ecological model (SEM).

## INDIVIDUAL FACTORS

a) Navigating the impact of unintended pregnancy

Among some adolescent teenagers discovering their pregnancy, they experienced significant psychological implications, triggering mental and emotional distress, heightening feelings of isolation, and exacerbating vulnerabilities. For some, the double burden of unintended pregnancy and living with HIV felt overwhelming. Awareness of different options regarding their sexual and reproductive healthcare after pregnancy awareness was burdensome and their mental and emotional capability to disclose this information was challenging.

*i) The double “burden” of pregnancy and living with HIV*

Fear of disclosing pregnancy as well as living with HIV to close friends and family led to mental health distress among adolescent girls. In some instances, teenage girls unknowingly became pregnant and concurrently contracted HIV, which intensified the impact of the news. According to a Healthcare Provider One:

*It becomes very difficult for the newly diagnosed and it is different compared to those who have been on treatment already because those who are on treatment, … continue with their treatment. For the newly diagnosed, maybe … she was shocked even by the fact that she is pregnant. It happens that one was just coming for family planning or came to remove the implant. It happens sometimes that the implant expired then she became pregnant, while she received the news of pregnancy she also received the news of being HIV-positive. So, you can see that she is facing two things at once.* [Healthcare Provider One]

For participants, such a double burden of major health challenges simultaneously resulted in acute shock and emotional distress, leading to mental unease.

ii) *Fear of disclosing pregnancy*

News of unplanned pregnancy and, for some, a new diagnosis of HIV, led to distress. For others, merely the news of an unplanned pregnancy was a shock leading to psychological distress.

*I did not notice it by myself but there were people [who] would make comments that "This one should be pregnant, why [is] she becoming fat?" and I would respond to them and say "No, I’m not pregnant"…I thought of buying a pregnancy test. I…did the test on my own. That’s when I discovered that I was pregnant… I was so fearful. I was afraid to disclose it at home.* [Postpartum and not living with HIV]

A participant, pregnant but not living with HIV said she was too afraid to notify her parents of her pregnancy and left them to notice this. Such thoughts emanated from fears of known or anticipated negative family reactions towards news of the unplanned pregnancy:

“*I was afraid [of] the fact that I disappointed my mother so I was afraid to talk about it at home”* [Postpartum, not living with HIV].

Another teenager mentioned:

*I was afraid and was very shocked* [to discover she was pregnant] … *afraid of what my parents would say… I thought that they would shout at me now that I had become pregnant.* [Pregnant and not living with HIV]

iii) *Options after pregnancy*

Most teenagers were unsure of what actions to take after realising that they were pregnant. Due to fear of disclosing pregnancy to parents as well as facing stigma from peers, some teenagers decided to terminate the pregnancy. However, some family members opposed the idea.

*When this child was told that she was pregnant, she decided that she was going to have an abortion … I screamed … ‘There are ten commandments that were written by God that you must not kill. Therefore, when you do the abortion you are killing’.* [Mother of Postpartum teenager, teenager not living with HIV]

This teenager opted to keep the pregnancy but have the baby adopted after birth, however, her parent also objected, agreeing to care for the baby:

*… if you take this baby and give [it] to someone we don’t know when the birthday of your child comes …. you will ask yourself questions as to what you have done. And all this will result* [in] *depression. So, I ask you to … accept that now you have a baby but this will be mine and not yours.* [Mother of Postpartum teenager, teenager not living with HIV]

Although the research method was unable to establish an association between pregnancy and negative mental health, the findings indicated that options after pregnancy created psychological burdens for some adolescent girls.

## INTERPERSONAL FACTORS

b.1.) Relationships in the home environment

Adolescents’ relationships with family members impact both their mental health and access to health care. Several factors such as a lack of family support, having a dysfunctional family, financial hardships at home, and the presence of domestic violence and abuse created a difficult environment for some participants, which likely led to mental distress among adolescents.

*i) Support from family/significant other*

Receiving family support was highlighted by several participants as a factor that would carry them through difficult moments in life. Unfortunately, the families of some adolescents did not support the unintended pregnancy:

*I did not know that I was pregnant … my mother noticed … She did not say anything at that time, she waited for me to go to school… When I came back from school she shouted at me and she called some other family members. Everybody was shouting at me … They tried to beat me … At the time I thought that no one wanted me in life because I am an orphan … I tried to commit suicide* [Post-partum and living with HIV].

The extreme action of wishing to commit suicide highlights the significant mental distress the teenager experienced from the family’s negative reaction towards unintended pregnancy.

Other families of adolescent girls were supportive. One participant, who was pregnant but not living with HIV, had battled with childhood trauma due to her father passing away when she was young. This participant was not living with her biological mother, who struggled with alcohol abuse. Her life challenges caused significant mental health distress. Fortunately, the participant’s stepmother, who she lived with, provided significant emotional support to her with words of encouragement:

*My stepmother is always there and encourage[s] me all the time… She told me that I should forget about things that happened in the past and always focus on the things of my school [school work].* [Pregnant and not living with HIV]

The encouraging words brought happiness to the participant, leading to a positive mental health impact. Another participant received support towards the pregnancy from her significant other.

*I… informed the father of the child … about my pregnancy and… he said he does not have a problem. He…supports me most of the time by buying a few items for me and I felt well. I did not have stress about what my child was going to wear because the father of the child was encouraging me and saying that he was going to take care of [the] child.* [Postpartum and not living with HIV]

Another participant also faced childhood trauma, losing her father to death at a very early life stage: “*The passing away of my father … it was very painful to me…I was thinking that I should just follow him as well* [Not pregnant and living with HIV]*”.* The participant received support and comforting words from relatives: *“My grandmother and grandfather were comforting me. They made me not to always think about my father’s death”*.

ii) *Dysfunctional family*

Dysfunctional family setup was a contributing factor to mental health distress among adolescents during their formative years. One adolescent participant described the challenges and adverse conditions in her home environment:

*My life was difficult because my mother left me when I was still young …My mother was addicted* [to] *drinking alcohol and …and she dumped me to my father. My father was not working during those times…*[he] *got some piece jobs as the time went by. From there, he decided to send me to his sister in rural areas.* [Pregnant and not living with HIV]

A difficult home environment, marked by feelings of early abandonment by a mother at a time when her presence was most needed, exposure to the mother’s alcohol addiction, fathers dying when they were young, and placement with other guardians was likely to contribute to the participants’ distress.

iii) *Financial hardships*

In some cases, financial hardships within families were significant contributing factors to mental distress. One participant, an adolescent who was neither pregnant nor HIV-positive, explained that life was fine until her father’s injuries disrupted the household income, resulting in considerable economic challenges:

*Life was all right, but my father got shot when I was in grade 7…He was doing security work…We were living rightfully except [for] this thing that happened … my father went to stay in the hospital for a long time…It was very difficult because he was the only person who was working at home.* [Not pregnant and not living with HIV].

iv) *Domestic violence and abuse*

Domestic violence and abuse were critical contributors to an adverse home environment, which, in turn, precipitated mental distress among adolescents. These harmed the home environment and impaired participants’ ability to concentrate on essential aspects such as education. These lead to considerable psychological stress among teenagers. “*I could not study well at home because there was always [a] lot of noise, people drinking, and constant fights. So, I could not do anything.”* [Postpartum and living with HIV]

A participant, who experienced challenges in school, having recently failed, highlighted the negative effects of poor domestic conditions:

*I am trying to focus but my family members are always in conflict among[st] each other … They drink almost every weekend and they open [play] loud music and I cannot find time to study my books well in that environment.* [Not pregnant and not living with HIV]

Persistent domestic conflicts and alcohol consumption, which sometimes escalated to physical violence, along with the playing of loud music in the home without regard for the needs of school- going adolescents, emerged as significant disruptive factors to educational progress and mental distress.

b.2.) Relationships with peers from school

Other interpersonal relations such as peer pressure, bullying, and stigma around teenage pregnancy in schools were also associated with psychological distress among teenage girls.

*i) Peer pressure*

Peer pressure and social comparisons were significant contributing factors to mental health challenges among teenagers. A healthcare provider explained as follows:

*I am also surrounded by teenagers in my house… we cannot run away from peer pressure as a contributing factor* [to mental distress]*. The more you want to compare yourself to someone else, the more stressful the situation becomes. A person ends up depressed because she wants something that another person is having while on her side she cannot afford.* [Healthcare provider Two]

Peer pressure could lead to feelings of inadequacy, causing low self-esteem and anxiety.

ii) *Bullying*

Bullying was another factor likely to result in mental distress among adolescents. Bullying was described as bullying (internet bullying through social platforms) or in-person bullying at schools., One of the program managers recounted:

*…I think adolescents face different challenges than what adults face. I heard that … adolescents … might be bullied and they might be stressed, they don’t want to go to school because they were bullied. So that also plays a… part in their mental health, in their development. I mean you have heard on the news how many suicides there have been because of bullying, because of teenagers and social media and the pressures they face. Now I think adolescents have different stressing things that adults don’t face like social media bullying and school bullying.* [Program Manager; FGD]

Sadly, in some cases, bullying, if not resolved timeously and appropriately, might result in extreme incidents, such as suicide among adolescents.

iii) *Stigma around teenage pregnancy in schools*

A further theme that emerged was participants being stigmatized by school peers for being pregnant. Some pregnant adolescent girls experienced gossip and, at times, mockery and remarks passed around by peers in the school environment:

*… they started gossip*[ing] *now that they knew about my pregnancy … they gossip whenever they talk among themselves. But they do not come to me… Sometime*[s] *whenever there is someone who get*[s] *drowsy in class, they will always make remarks that “There is someone pregnant among us”. …“They make me feel sad*” [Pregnant and not living with HIV]

Gossip and stigmatization from peers caused deep pain and distress to participants interviewed and she began to cry while narrating her experience. Peer stigmatisation can be a powerful factor negatively impacting mental health and well-being.

## ORGANISATIONAL FACTORS

c) Barriers around access to services at facility level

*i) Contraceptive services*

Adolescents recounted receiving information about contraceptives from various sources. For some, clinics, schools, and youth centres provided information:

*I found it from the clinic which was there in the village. I started using them in 2018 because our school is close to the clinic. There … nurses [would] come out from the clinic and call us on the school lines and talk about the family planning methods.* [Postpartum and not living with HIV]

For others, the information about contraceptives was acquired from friends and family.

*The information about family planning methods I found it from my friend who I trust. I started telling her about things that were happening … [at] home and then she said "My friend you must go to the clinic to protect yourself as you know how the situation is, people arrive in your home drunk and some of them keep on touching you and I don’t know what will happen. So it is better for you to protect yourself".* [Postpartum and living with HIV]

A healthcare provider shared her views and observations concerning contraceptive information and awareness among teenagers:

*Adolescents who come here for the first time* [are] *mostly accompanied by their parents. If …not…when they come here, they come with the information that their parents have already told them about which method they should choose. We also play our role in explaining to them about different methods that are available so that they can be able to decide on their own as they don’t normally need parental consent. You would explain to them about the Noristerat for the two months or Petogen which is taken for 3 months. There is also an implant for three years and the IUD for 5 years. They also have their own choice to make… at least they must know a variety of methods that are available so that one cannot just only choose the method that the mother has recommended.* [Healthcare Provider One]

Healthcare providers play a pivotal role in sharing information about the different family planning methods with adolescents, increasing their knowledge and awareness of contraceptives.

However, some misinformation and misconceptions on contraceptive use became a barrier for some using these methods and exposed them to the risk of pregnancy and STIs:

*I guess I was scared as I heard some negative stories about them … One of my friends who was in class with me said … “I think the injectable makes you fat”* [Pregnant and not living with HIV]

A healthcare provider too, highlighted perceived myths around the use of contraceptive methods.

*Sometimes they come with myths that they hear from their communities about what the family planning does. For example, [like] if one uses family planning she may end up not being able to conceive in future. Maybe someone comes already with the information from her mother in a way that she must take the Noristerat because Petogen is for older people. You would find that when they come here, that information already sits in a child’s mind. Again, they [are] present[ed] with a myth that family planning disturbs their periods [menstruation] process in a way that they become irregular. As a result you noticed that some of them end up stopping taking or using family planning.* [Healthcare Provider One]

Increasing the awareness and the sources of correct information about contraceptives can assist teenagers in making better-informed decisions about their sexual and reproductive healthcare.

ii) *Judgement from healthcare workers*

Another element mentioned by one of the healthcare providers impeding adolescent access to some healthcare services was judgment by healthcare providers, which should cease:

*I feel like there is still a lot more that needs to be done because other diseases come up and pass but the stigma that is with HIV is still difficult to remove from other people. So, we* [healthcare providers] *must try to talk to the clients as healthcare providers and not judge a child and say, “No, a child at the age [of] 18, what were you doing?” and things like that. A child must be comfortable to talk with us about everything that she feels …. This will help a lot so that clients will not default.* [Healthcare Provider Two]

A consequence of such judgment from healthcare workers is teenage resistance to accessing these services. The same healthcare provider highlighted that some patients living with HIV would rather default on their HIV medication and treatment than engage with judgmental healthcare providers, thus, acting as a barrier to access to services at the facility level.

iii) *Unawareness of mental health policies by youth service providers*

Some providers were familiar with HIV guidelines but lacked knowledge of mental health guidelines:

*I don’t know about guidelines at all, child and adolescent mental health policy, currently being rolled out at the provincial level … this is the first time I hear about it … I know only HIV guidelines.* [Healthcare Provider One]

The majority of youth healthcare service providers interviewed were either unaware of mental health policies, including broader child and adolescent policies, or had only heard about them without having read them:

*I am not aware of* [the] *new adolescent mental health policy. I don’t deal much with mental health issues in depth … I remember that there were earlier policies* [Healthcare Provider Four]

Healthcare providers demonstrated a lack of adequate specific knowledge about mental health guidelines, tailored for youth. This knowledge gap poses a significant barrier to their ability to adequately provide mental health care to adolescents experiencing mental health challenges. Without a comprehensive understanding of these guidelines, the detection, diagnosis, and treatment of mental health conditions teenagers will experience exclusion from mental health services.

iv) *Inadequate healthcare facilities for adolescents*

An organisational barrier highlighted by participants as limiting the accessibility of mental health services was the lack of sufficient infrastructure. While the need for such mental health services exists the available infrastructure to provide these is limited. A district manager explained:

*… the infrastructure that we have in primary care is hopelessly inadequate, buildings are too small, we have become comprehensive, and we take on adolescent care and … mental health, we operate in the same building … but the building space does not allow* [for mental health service provision] *…* [District Manager; FGD]

She continued to emphasise that the limited infrastructure at the healthcare facility compromised the provision of one service (mental health services) at the expense of another (provision of medicine). However, providing medicine to adolescents alone is insufficient, counselling and therapy sessions are needed.

*v) Healthcare professional shortages*

Having a shortage of healthcare professionals was a further organisational barrier limiting adolescents’ access to mental health services. There are too few professional psychologists available to provide adequate counselling sessions:

*We have been introduced to the psychologists. I just can’t remember how many we have in the whole City [of Cape Town]. Is it maybe 4? So how do you introduce 4 people into a service where there is like 50% of people that* [need them] *…Who do you refer to them? Who do you take?* [District Manager; FGD]

Decision-making on who should receive such counselling from professional psychologists becomes difficult. Inevitably some adolescents will not receive counselling sessions, increasing their risk of anxiety and depression

## COMMUNITY FACTORS

d) Social challenges

*i) Social stigma around teenage pregnancy*

Societal judgment has mental health implications for pregnant teenagers. A participant recalled how she was already treated differently by community members when she was living with HIV but that this was exacerbated when she became pregnant.

*They* [the community members] *did not make me feel happy…During the time I became pregnant, there were some people … judging me that "Yho!* [a mocking expression of surprise, saying] *we knew that she was going to be pregnant and…*[she] *is not going to continue with school anymore".* [Postpartum and living with HIV]

Such judgemental attitudes from community members can lead to negative mental health outcomes for participants.

ii) *Substance abuse*

The abuse of substances such as alcohol, cannabis, and other harmful drugs, is heightened as a community hazard for adolescents in communities:

*It has a lot of contribution because some of them* [teenagers] *would like to experience everything when they reach* [the] *teenage stage and they do not want to remain behind… A certain group will be against another group and there are those infighting at school among those grouping*[s] *and they use the substance and a person end*[s] *up depending on that substance. This all leads to someone among them becoming mentally disturbed.* [Healthcare Provider Two]

Heavy dependency on these substances and conflicts about them within schools can lead to mental health deterioration resulting in mood disorders, anxiety, and depression among teenagers.

iii) *Unsafe communities*

Lack of safety in communities is an important factor that can lead to fear and anxiety among adolescent girls. Acts of sexual violence, particularly rape of children and women were major concerns causing a sense of insecurity in some communities. One of the participants narrated the following regarding the issue of safety within their community:

*Sometimes you’d hear …[about] children who got raped and some other people who are suffering in the communities … People who are raping women [and] children they must be caught and put in prison so that we can be safe in the communities.* [Postpartum and not living with HIV]

Living in unsafe communities also a potential site of sexual violence, can lead to significant mental distress, particularly in adolescent girls.

## POLICY LEVEL ISSUES - PROVISION OF MENTAL HEALTH SERVICES

*e) Lack of PACK guidelines for adolescents*

Nationally recognised policy guidelines for mental health are important. The Practical Approach to Care Kit (PACK) was developed as a comprehensive set of guidelines with the primary objective of guiding clinical decision-making during patient consultations in South Africa (Cornick et al., 2018). These guidelines are to ensure that care aligns with policy and delivers integrated, comprehensive primary health care.

However, while well-developed versions exist for children under 13 (PACK Child) and adults (PACK Adult), currently there are no finalised PACK guidelines for adolescents. One of the district managers described the situation, “*… we don’t have the adolescent PACK guidelines but it is actually coming soon. So that actually focuses … on adolescents…”,* [District Manager; FGD.

Such lack of specific adolescent PACK guidelines increases the vulnerabilities of adolescents to misdiagnoses and inadequate treatment and care for health conditions, particularly mental health issues.

### Discussion and conclusion

The findings of this study reveal a complex interplay of factors contributing to adolescent mental health challenges, exacerbated by pregnancy and HIV. Influences range from individual, family, organisational, community, and health systems domains. At an individual level, unintended pregnancy, particularly when coupled with living with HIV, can lead to mental distress emanating from fear of disclosure, shock, and pervasive stigma around unintended pregnancy. This double burden increases feelings of isolation, anxiety, and depression among adolescent girls. This echoes findings elsewhere that unintended pregnancies disproportionately impact adolescent girls, especially when intersecting with socio-economic vulnerabilities and HIV (Toska et al., 2020; Erasmus, Knight & Dutton, 2020; WHO, 2022; Mkhize et al., 2024).

Within the interpersonal, family domain, adverse home environments marked by a lack of family support, dysfunctional family dynamics, financial hardships, and domestic violence and abuse emerge as critical determinants of poor mental health. The absence of consistent emotional support and open communication not only deepens feelings of isolation and despair but may also negatively influence future sexual and reproductive decisions. These findings are consistent with evidence that stable, supportive, safe, and conducive home environments are crucial for fostering adolescent resilience against mental distress and other life challenges (Mkhize et al., 2024; Patton et al., 2022). Bullying at school and elsewhere is highly prevalent and needs to be addressed as a major stressor for adolescents (Källmén & Hallgren, 2021). HIV emerged as an exacerbating factor for example in one participant’s comments on having already been gossiped about by community members while living with HIV before becoming pregnant. This worsened after she became pregnant, leading to much stress. Healthcare providers also commented on the additional shock adolescent girls experience when, in addition to the diagnosis of HIV, they were told they were pregnant. In these cases, the negative feelings were extreme to the extent that one of the adolescents wished to commit suicide. This shows that the burden of falling pregnant and concurrent HIV acquisition can be a big trigger for potential mental health issues. This may have lifelong implications for the adolescents’ mental health. Established relationships in schools and communities have an impact on mental health, and maintaining stable interpersonal relationships can significantly contribute to mental well-being among adolescents.

At an organisational level, certain barriers to health services, including misinformation about contraceptives from peers or their communities, judgment from some healthcare providers, the lack of mental health information from some healthcare professionals, inadequate infrastructure, and a shortage of professional psychologists can increase adolescents’ vulnerability to mental health issues.

At the community level, the pervasive social stigma surrounding teenage pregnancy significantly contributes to adverse mental health outcomes, exacerbating feelings of shame and low self-esteem (Erasmus et al., 2020). In addition, the prevalence of substance abuse of alcohol, cannabis, and other drugs in these communities is linked to increased rates of mood disorders, anxiety, and depression among teenagers (Shisana et al., 2024). Moreover, living in unsafe communities where incidents of violence including sexual violence are common further intensifies feelings of insecurity and anxiety, particularly among adolescent girls. This underscores the urgent need for comprehensive community-based interventions and enhanced legal protections (Toska et al., 2020; Mokitimi et al., 2019).

Within the policy level domain, significant gaps exist in the design and delivery of mental health services tailored for adolescents. This is highlighted by the lack of adolescent PACK guidelines. Using adult PACK guidelines for adolescents can lead to misdiagnosis as mental health issues amongst adults mostly differ from adolescents. Thus, adult PACK guidelines fail to meet the specific needs of this group, necessitating targeted interventions and specialized training for healthcare providers. Even if adolescent PACK guidelines are developed and implemented, if there is limited awareness of these guidelines and inadequate training of some healthcare providers, challenges to meeting adolescents’ mental health needs will remain (Smith & Nkosi, 2020; Mhlongo et al., 2019; Petersen et al., 2020).

The results showed that adolescents who were pregnant/postpartum and infected with HIV experienced more severe mental health issues than other participants (those who were not pregnant and/or infected with HIV). Lack of family support during pregnancy, created a greater adverse home environment, contributing to mental health distress among pregnant adolescents. These findings are supported by other studies such as Mosaya et al. (2022) where they found a higher prevalence of depression among pregnant women compared to non-pregnant adolescents. Similarly, a study in rural South Africa found that pregnant and post-partum teenagers experienced various mental health challenges due to stigmatisation, lack of support from family and friends, parenting demands, and poor performance in school (Muthelo et al., 2024). Additionally, a study in Uganda (Kesande et al., 2023) also observed a higher prevalence of psychological distress amongst pregnant young women living with HIV compared to non-pregnant women living with HIV, findings that are similar and support our findings. Hence, there is a need for comprehensive mental health intentions and other strategies for all adolescent girls, particularly those who are pregnant and/or are infected with HIV.

Overall, the findings underscore the urgent need for more comprehensive approaches that simultaneously address individual vulnerabilities, strengthen interpersonal relationships, manoeuvre around social challenges, and resolve organisational and policy-level barriers to mental health services. Interventions tailored for adolescent girls are essential for mitigating the intersectoral factors contributing to mental health challenges that impact adolescent girls and for advancing their access to appropriate healthcare services.

## Data Availability

Data cannot shared publicly because it was collected for human participants who were promised anonymity. However, data can be made available upon request.

## Acknowledgments

We acknowledge the support of all participants who took part in this study. These include adolescent girls, the parents of adolescent girls, healthcare providers in health facilities, and senior managers. We are grateful for their time and for the rich information, they shared with us. We also acknowledge our data collector Ntobeko Nywagi, who worked so hard to find adolescent girl participants and interview them. Lastly, this work is based on the research supported in part by the National Research Foundation of South Africa (Grant Numbers: 11682 & 82769). The authors would also like to acknowledge the support of the South African Medical Research Council, through the Health Services to Systems Extra Mural Research Unit. Any opinion, findings, conclusion, or recommendation expressed in this material is that of the authors and not the funders.

## Author’s contributions

All authors contributed substantially to this work. HT and ML conceptualised the study, designed the research methodology, and managed data collection. HT, TS, DC, and ML were involved in data analysis and manuscript drafting. All the authors read and commented on all the drafts and the final manuscript.

